# SARS-CoV-2 infections and hospitalizations among immigrants in Norway – significance of occupation, household crowding, education, household income and medical risk. A nationwide register study

**DOI:** 10.1101/2021.05.04.21256575

**Authors:** Angela S Labberton, Anna Godøy, Ingeborg Hess Elgersma, Bjørn Heine Strand, Kjetil Telle, Trude Arnesen, Karin Maria Nygård, Thor Indseth

## Abstract

**Background:** As in other countries, the coronavirus disease 2019 (COVID-19) pandemic has affected Norway’s immigrant population disproportionately with significantly higher infection rates and hospitalizations. The reasons for this are uncertain.

**Methods:** Through the national emergency preparedness register, BeredtC19, we have studied laboratory-confirmed infections with SARS-CoV-2 and related hospitalizations in the entire Norwegian population, by birth-country background for the period 15 June 2020 – 31 March 2021, excluding the first-wave due to limited test capacity and restrictive test criteria. Straightforward linkage of individual-level data allowed adjustment for demographics, socioeconomic factors (occupation, household crowding, education, and household income), and underlying medical risk for severe COVID-19 in regression models.

**Results:** The sample comprised 5.49 million persons, of which 0.91 million born outside of Norway, 82 532 confirmed cases, and 3088 hospitalizations. Confirmed infections in this period (per 100 000): foreign-born 3140, Norwegian-born with foreign-born parents 4799, Norwegian-born with Norwegian-born parent(s) 1011. Hospitalizations (per 100 000): foreign-born 147, Norwegian-born with foreign-born parents 47, Norwegian-born with Norwegian-born parent(s) 37. The addition of socioeconomic and medical factors to the base-model (age, sex, municipality of residence) attenuated excess infection rates by 12.0% and hospitalizations by 3.8% among foreign-born, and 10.9% and 46.2% respectively among Norwegian-born with foreign parents, compared to Norwegian-born with Norwegian-born parent(s).

**Conclusion:** There were large differences in infection rates and hospitalizations by country background, and these do not appear to be fully explained by socioeconomic and medical factors. Our results may have implications for health policy, including the targeting of mitigation strategies.

## INTRODUCTION

This short communication is based on a Norwegian-language report on the implications of socioeconomic and medical risk factors on coronavirus disease 2019 (COVID-19) among immigrant groups, produced by the Norwegian Institute of Public Health (NIPH) in April 2021 (1). It is important to disseminate more widely new knowledge that may help inform health policy, including targeting mitigation strategies toward the most affected groups, to help prevent hospitalizations and deaths from COVID-19.

Norway has thus far had relatively low rates of COVID-19. Since the first case in February 2020 and until 18th April 2021, there have been, per 100 000 population: 2002 confirmed cases, 74.4 hospitalizations, and 13.5 deaths. Still, the immigrant population has been hit disproportionately (2), as is also documented in several other countries (3-6). Country background may be related to several factors affecting the risk of COVID-19 infection and/or hospitalization, such as socioeconomic status, occupation, and health disparities. However, studies of how all these factors affect COVID-19 in immigrant populations are lacking. In the United Kingdom (UK) it appears that only part of the elevated risk of infection (4), hospitalisation (5), and death (7, 8) can be explained by socioeconomic factors (varying definitions). In Norway, foreign-born persons more often live in the large cities, particularly Oslo, where infection rates have been high. Additionally, they more often live in overcrowded housing and on average have lower income compared to Norwegian-born persons. We have investigated whether such observable characteristics explain differences in infection and hospitalization rates between persons with different country backgrounds.

## METHODS

The BeredtC19 Register is a national emergency preparedness register established during the COVID-19 pandemic (https://www.fhi.no/en/id/infectious-diseases/coronavirus/emergency-preparedness-register-for-covid-19/). BeredtC19 contains individual-level data, covering the entire Norwegian population, linked via the unique, personal identifier given to all Norwegian residents at birth or upon immigration. The BeredtC19 data used for this study originated from the Norwegian Surveillance System for Communicable Diseases and laboratory database (all polymerase chain reaction (PCR) tests with test results for SARS-COV-2), Norwegian Patient Registry and Norwegian Registry for Primary Health Care (hospital admissions, medical risk group), National Population Register (demographics, municipality of residence), Employer and Employee Register (occupation), and Statistics Norway (household crowding, education, household income). See supplemental material for full methods and variable definitions. Briefly, household crowding is a pre-defined indicator variable and is present if the dwelling har fewer rooms than number of residents (or one person in a one room dwelling), *and* the internal area is <25 m2 per person. Highest education, available until 2019, was categorized into ‘Below upper secondary’; ‘Upper secondary/vocational’; ‘University/college, short’; ‘University/college, long’; and ‘Undisclosed/no education’. Persons <26 years old (N=1 606 768), may not yet have completed their education, and were coded into a separate category. Household income was recorded as annual household income after tax, divided by number of consumptions units (EU-scale) in the household and categorized in deciles. Medical risk groups are a defined set of 14 diagnoses or health conditions identified by the NIPH to convey higher risk of severe COVID-19 (requiring hospitalization), these groups are categorized as present or not. Persons with missing data for any explanatory variable were coded into a separate residual category for that variable and included in analyses, thus keeping the same sample for all analyses.

We studied two outcomes: laboratory-confirmed (PCR) infection with SARS-CoV-2, and hospitalization with COVID-19. We compared residents firstly as three groups: i) Foreign-born, ii) Norwegian-born with foreign-born parents, iii) Norwegian born with ≥1 Norwegian-born parent; and also analysed foreign-born persons from the 25 birth-countries with >10 000 residents in Norway compared to all Norwegian-born persons (regardless of parental birth-country). Linear regressions were used to estimate the following models: (1) Unadjusted; (2) Age, sex; (3) Age, sex, municipality of residence (Base-model); (4) Base-model + occupation; (5) Base-model + household crowding; (6) Base-model + education; (7) Base-model + household income; (8) Base-model + medical risk for hospitalization with COVID-19; and (9) All factors. We studied the period 15 June 2020 – 31 March 2021, excluding the first-wave due to limited test capacity and restrictive test criteria.

## RESULTS

The sample comprised 5.49 million persons, of which 0.91 million born outside of Norway, 82 532 confirmed cases, and 3088 hospitalizations (Table 1). Norwegian-born to foreign parents (N=199 518) had highest infection rates, and higher hospitalizations than Norwegian-born to Norwegian-born parent(s). Persons born outside of Norway had highest rates of hospitalizations. There was large variation between different country backgrounds. Infection rates were highest among persons born in Pakistan, Somalia, and Iraq; and lowest for China, Germany, and Denmark. Hospitalizations were highest for Pakistan, Iraq and Turkey; and lowest for USA, Lithuania and Latvia.

**Table 1.** Numbers of confirmed cases with SARS-COV-2 and related hospitalizations by country of birth. Estimates from full model shown (age, sex, municipality of residence, occupation, household crowding, education, household income, medical risk).

| Country of birth | Number of cases | Cases per 100 000 | Number of hospitalizations | Hospitalizations per 100 000 | N | Estimates from model adjusted for all factors (robust standard error) |  |
| --- | --- | --- | --- | --- | --- | --- | --- |
|  |  |  |  |  |  | Cases per 100 000 | Hospitalizations per 100 000 |
| **Norway** | 53890 | 1175 | 1741 | 37 | 4582626 | 1276 (5.60) | 39 (1.00) |
| - with Norwegian-born parent(s) | 44315 | 1011 | 1647 | 37 | 4383108 | 1153 (5.41) | 39 (0.995) |
| - with foreign-born parents | 9575 | 4799 | 94 | 47 | 199518 | 3975 (48.36) | 53 (5.34) |
| **Outside of Norway** | 28642 | 3140 | 1347 | 147 | 912043 | 2636 (19.07) | 139 (4.21) |
| Afghanistan | 1118 | 6407 | 59 | 338 | 17449 | 5303 (184.60) | 335 (44.00) |
| BA-XK-HR-ME-RS-SI* | 1613 | 4087 | 95 | 240 | 39462 | 3389 (99.00) | 223 (24.60) |
| China | 114 | 855 | <5 | 30 | 13323 | 392 (80.20) | 35 (15.10) |
| Denmark | 295 | 1132 | 15 | 57 | 26051 | 1134 (65.50) | 35 (14.90) |
| Eritrea | 1272 | 5669 | 36 | 160 | 22437 | 5011 (154.50) | 177 (27.00) |
| Ethiopia | 508 | 4712 | 30 | 278 | 10780 | 3810 (203.10) | 267 (50.70) |
| Germany | 325 | 1078 | 14 | 46 | 30127 | 1109 (59.60) | 46 (12.50) |
| India | 433 | 2465 | 33 | 187 | 17563 | 1918 (117.40) | 175 (32.90) |
| Iran | 754 | 3932 | 50 | 260 | 19174 | 3183 (139.70) | 235 (36.80) |
| Iraq | 1711 | 7397 | 104 | 449 | 23130 | 6319 (171.00) | 421 (43.90) |
| Latvia | 153 | 1293 | <5 | <42 | 11829 | 1130 (104.70) | 43 (15.00) |
| Lithuania | 623 | 1444 | 7 | 16 | 43139 | 1246 (59.30) | 34 (6.90) |
| Pakistan | 2042 | 9173 | 200 | 898 | 22259 | 7562 (192.80) | 819 (63.00) |
| Philippines | 490 | 1855 | 24 | 90 | 26412 | 1416 (83.10) | 94 (18.70) |
| Poland | 3163 | 2954 | 55 | 51 | 107054 | 2534 (52.20) | 52 (7.40) |
| Romania | 409 | 2487 | 13 | 79 | 16442 | 2168 (122.00) | 86 (22.00) |
| Russia | 746 | 3948 | 44 | 232 | 18894 | 3693 (140.50) | 241 (35.10) |
| Somalia | 2395 | 8477 | 108 | 382 | 28250 | 7056 (166.20) | 353 (36.70) |
| Sweden | 877 | 1738 | 15 | 29 | 50449 | 1382 (58.30) | 13 (7.90) |
| Syria | 1415 | 4219 | 56 | 166 | 33535 | 3569 (110.00) | 191 (22.50) |
| Thailand | 350 | 1470 | 18 | 75 | 23797 | 1329 (78.70) | 97 (18.00) |
| Turkey | 705 | 5067 | 56 | 402 | 13913 | 4246 (184.70) | 376 (53.70) |
| United Kingdom | 273 | 1237 | 10 | 45 | 22056 | 1179 (74.60) | 34 (14.40) |
| United States of America | 251 | 1185 | <5 | <24 | 21179 | 1108 (74.60) | 6 (8.30) |
| Vietnam | 309 | 2086 | 34 | 229 | 14813 | 1318 (117.10) | 193 (39.30) |

\*BA-XK-HR-ME-RS-SI: Bosnia and Herzegovina, Kosovo, Croatia, Montenegro, Serbia and Slovenia.

There was large variation in the distribution of the socioeconomic factors and medical risk factors included by country of birth (Fig. 1). Foreign-born persons were more likely to live in crowded housing, have lower education and household income. Except for Pakistan, persons born outside of Norway were less likely to have a medical diagnosis associated with severe COVID-19. For some variables, we only had access to data until 2018 (household crowding, education) or 2019 (household income). Missing data was therefore higher among persons born outside of Norway, who may have immigrated after these dates. Missing: household crowding (N=405 642), 16.1% foreign-born vs. 5.7% Norwegian-born; household income (N=242 299), 10.8% vs. 3.1%; and for education (N=198 781), 19.1% vs. 0.5% had no or undeclared education (ages >25).

**Fig. 1.**
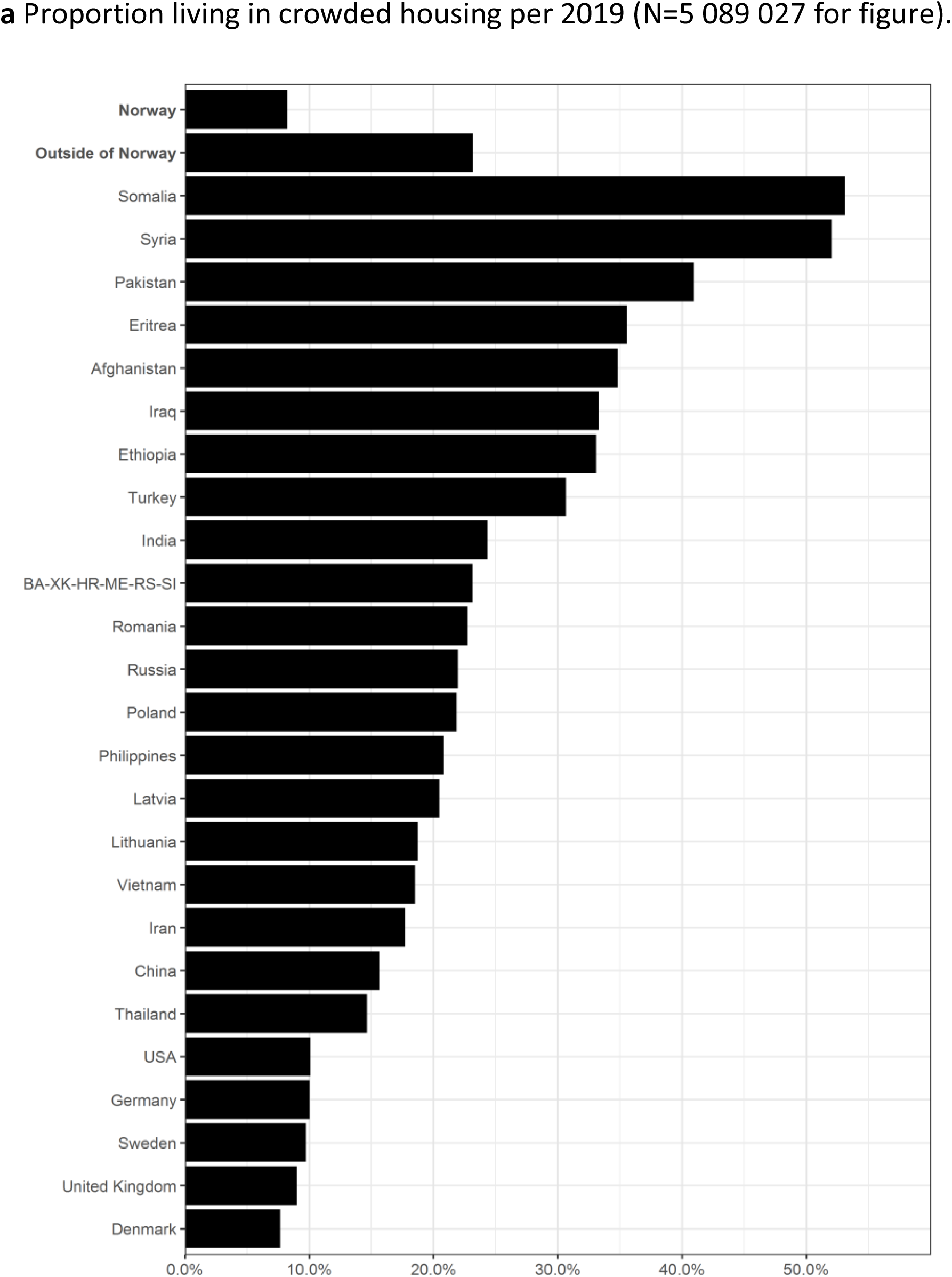
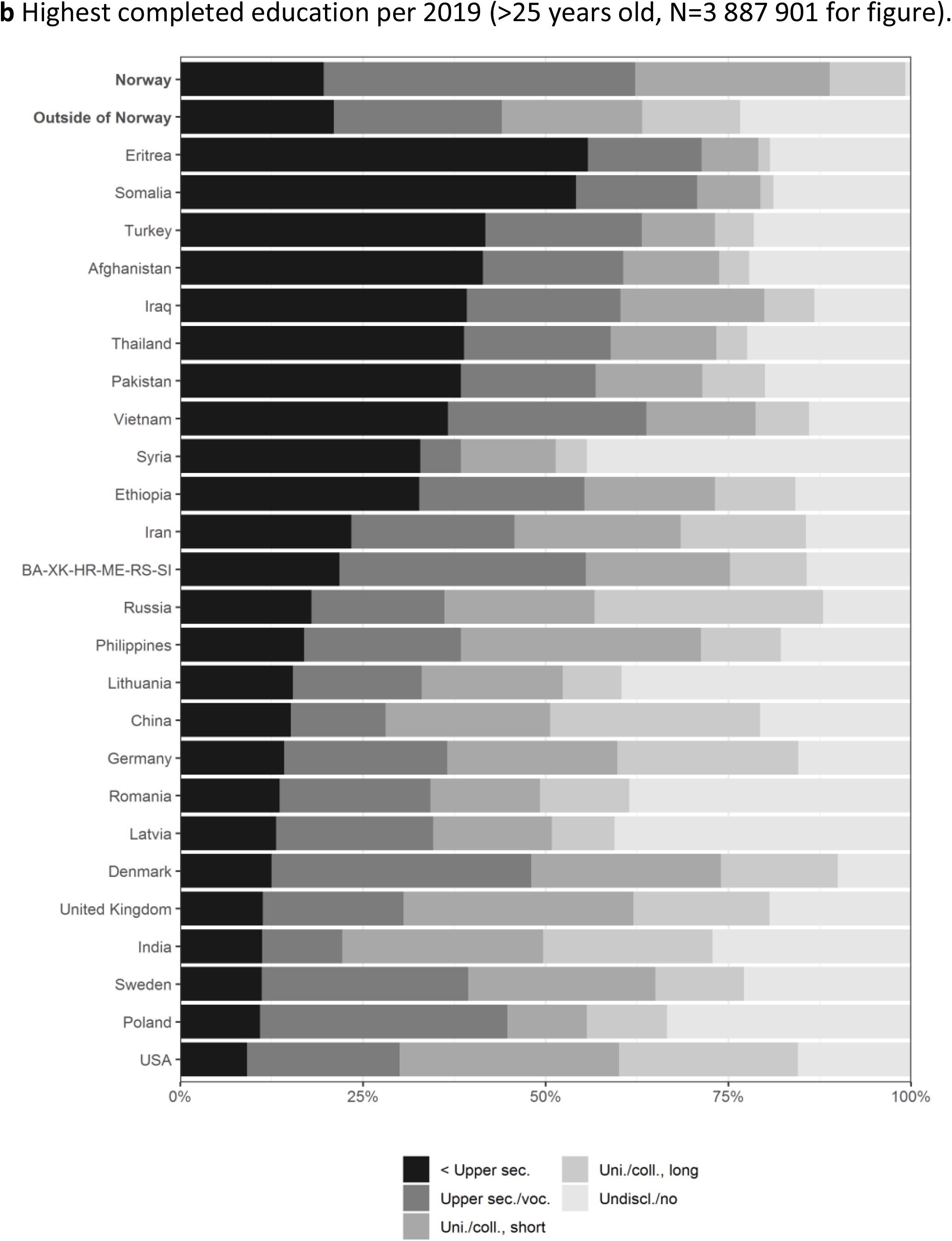
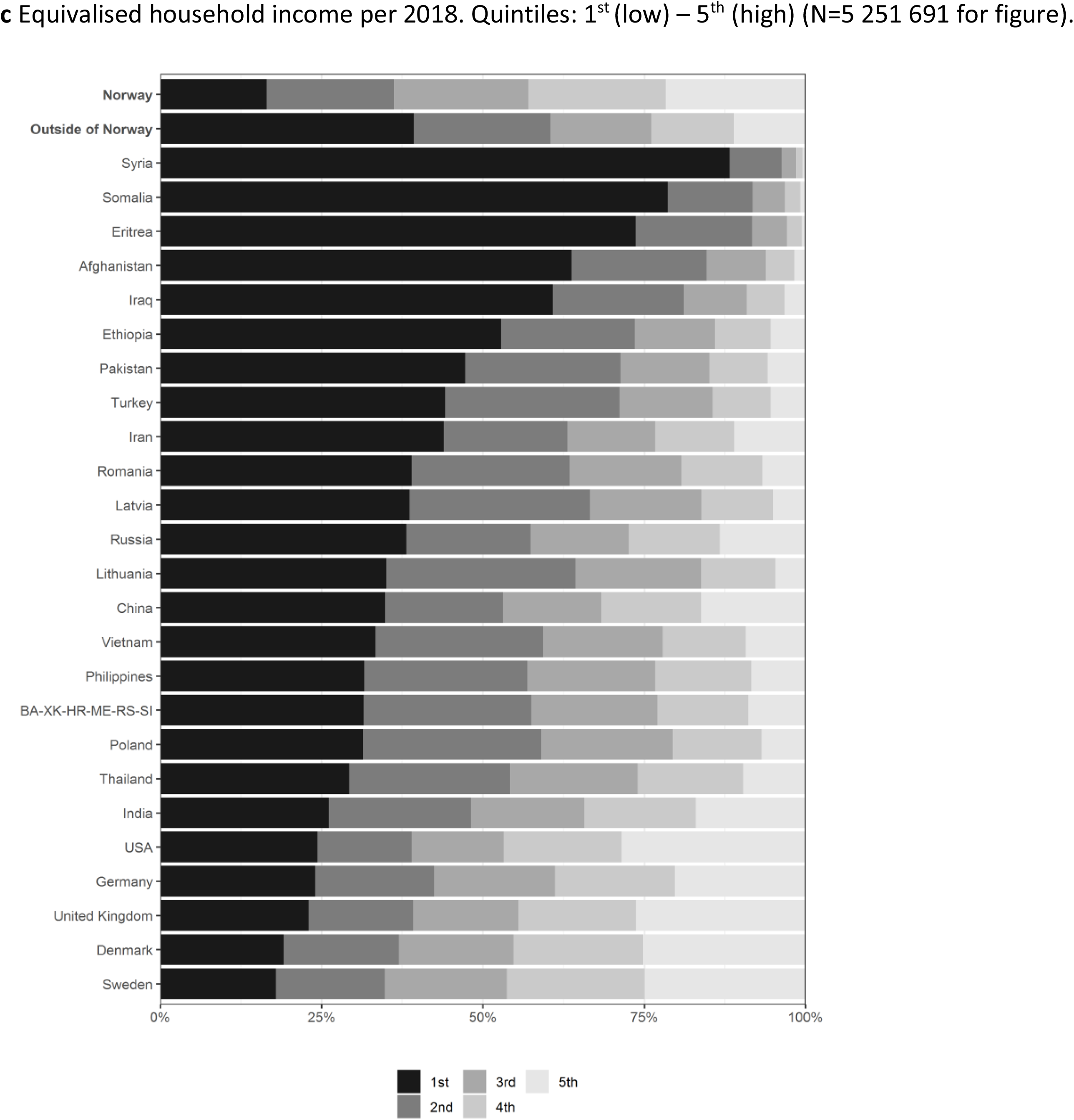
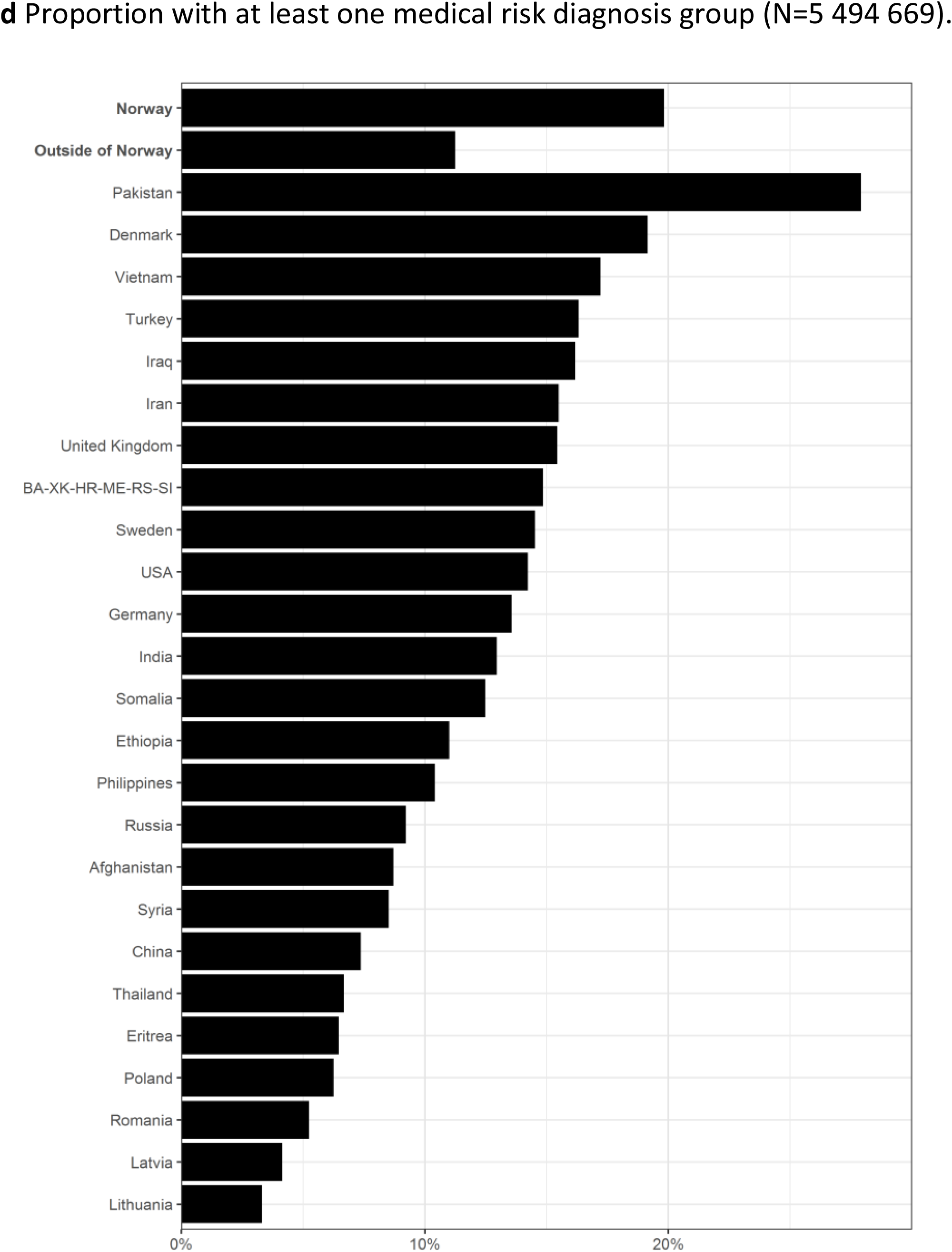
Distribution of socioeconomic factors and medical risk by country of birth. **a** Proportion living in crowded housing per 2019. **b** Highest completed education per 2019. **c** Equivalised household income (quintiles) per 2018. **d** Proportion with at least one medical risk diagnosis group.

All factors studied were each associated with infection and/or hospitalization with COVID-19 among both Norwegian-born and foreign-born persons. When the socioeconomic and medical factors were added to the base-model (age, sex, municipality of residence), excess infection rates were attenuated by 12.0% and hospitalizations by 3.8% among foreign-born, and 10.9% and 46.2% respectively among Norwegian-born with foreign parents, compared to Norwegian-born with Norwegian-born parent(s) (Table 2). In total, the full model attenuated excess infections compared to Norwegian-born with Norwegian-born parent(s) by 30.3% among foreign-born, and by 25.5% among Norwegian-born with foreign-born parent(s), compared to unadjusted estimates.

**Table 2.** Confirmed cases with SARS-COV-2 and related hospitalizations. Adjustments for age, sex, municipality of residence, occupation, household crowding, education, household income, and medical risk group for severe disease. Numbers per 100 000 (robust standard errors in parentheses).

|  | (1)<br>Unadjusted | (2)<br>Age, sex | (3)<br>Base-model:<br>Age, sex,<br>municipality<br>of residence | (4)<br>Base-model<br>+ occupation | (5)<br>Base-model<br>+ household<br>crowding | (6)<br>Base-model<br>+ education | (7)<br>Base-model<br>+ household<br>income | (8)<br>Base-model<br>+ medical<br>risk group | (9)<br>All factors |
| --- | --- | --- | --- | --- | --- | --- | --- | --- | --- |
| **Country of birth** |  |  |  |  |  |  |  |  |  |
| **Cases** |  |  |  |  |  |  |  |  |  |
| A) Norway, Norwegian-born parent(s) | 1011 (4.78) | 1038 (4.91) | 1107 (5.14) | 1114 (5.17) | 1138 (5.23) | 1106 (5.26) | 1129 (5.24) | 1107 (5.14) | 1153 (5.41) |
| B) Norway, foreign-born parents | 4797 (47.83) | 4839 (49.34) | 4273 (48.63) | 4263 (48.61) | 4054 (48.42) | 4255 (48.63) | 4147 (48.51) | 4273 (48.63) | 3975 (48.36) |
| C) Foreign-born | 3140 (18.26) | 3002 (18.35) | 2793 (18.21) | 2764 (18.26) | 2691 (18.15) | 2801 (19.09) | 2716 (18.38) | 2792 (18.20) | 2636 (19.07) |
| Percentage change in excess cases from base-model, group B) vs. A) |  |  | Ref. | -0.54 | -7.90 | -0.54 | -4.67 | 0.00 | -10.87 |
| Percentage change in excess cases from base-model, group C) vs. A) |  |  | Ref. | -2.14 | -7.89 | 0.53 | -5.87 | -0.06 | -12.04 |
| **Hospitalizations** |  |  |  |  |  |  |  |  |  |
| A) Norway, Norwegian-born parent(s) | 38 (0.93) | 34 (0.87) | 38 (0.95) | 39 (0.97) | 38 (0.96) | 38 (0.97) | 39 (0.97) | 38 (0.94) | 39 (0.995) |
| B) Norway, foreign-born parents | 47 (4.86) | 88 (5.28) | 64 (5.23) | 63 (5.22) | 58 (5.31) | 62 (5.23) | 54 (5.27) | 64 (5.23) | 53 (5.34) |
| C) Foreign-born | 148 (4.02) | 157 (4.17) | 142 (4.06) | 137 (4.05) | 142 (4.09) | 143 (4.22) | 139 (4.06) | 142 (4.05) | 139 (4.21) |
| Percentage change in excess hospitalizations from base-model, group B) vs. A) |  |  | Ref. | -7.69 | -23.08 | -7.69 | -42.31 | 0.00 | -46.15 |
| Percentage change in excess hospitalizations from base-model, group C) vs. A) |  |  | Ref. | -5.77 | 0.00 | 0.96 | -3.85 | 0.00 | -3.85 |

The same models were estimated for each of the 25 specified birth-countries (reference category: Norwegian-born), see Table 1 (last columns) and Fig. 2. While there was some variation in the relative significance of the different covariates between countries, no model changed estimates substantially (Fig. 2). Estimates for hospitalizations were less certain due to small numbers, but no adjustments had large effects (Fig. 2b).

**Fig. 2.**
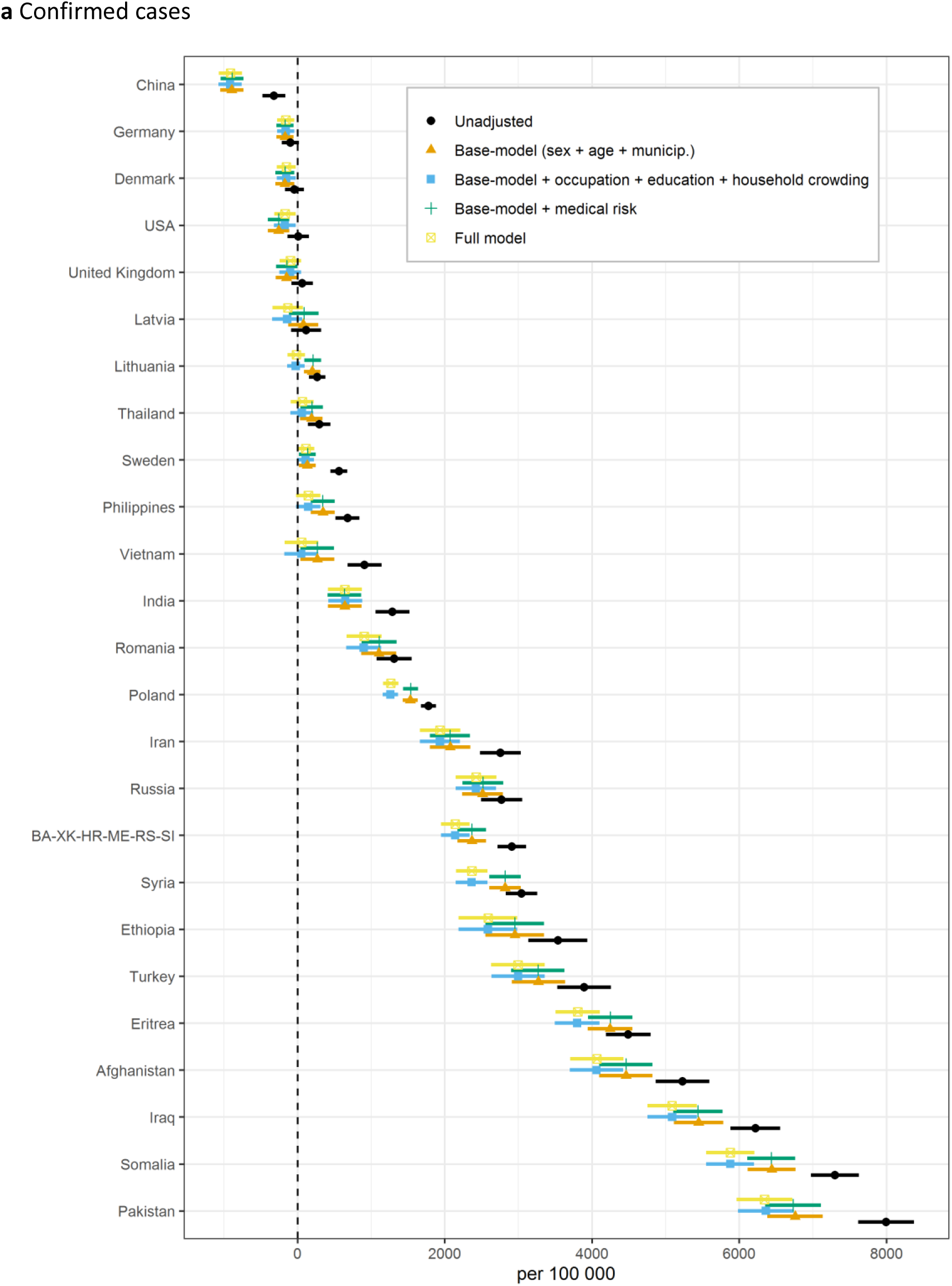
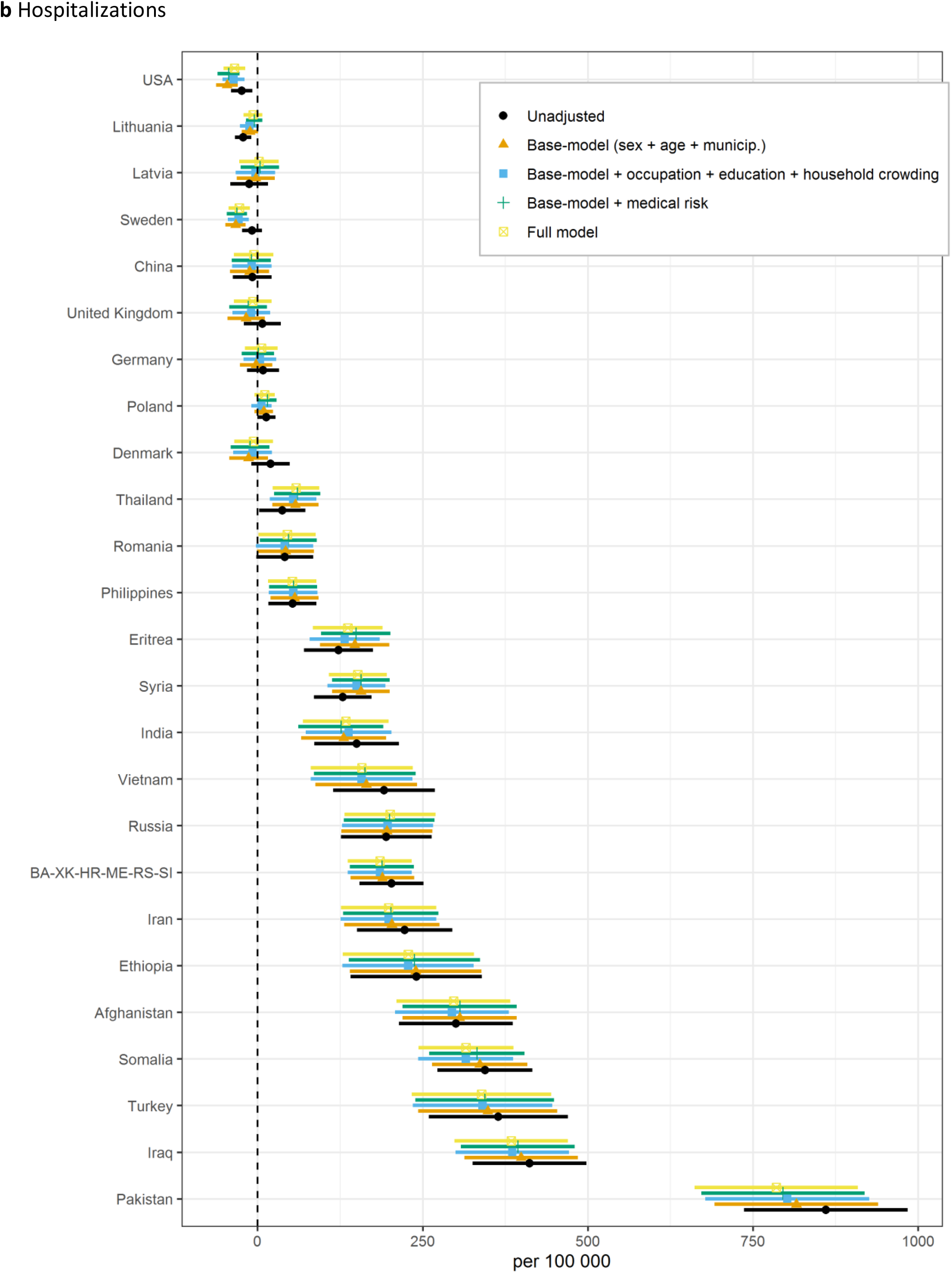
**a** Confirmed SARS-CoV-2 cases and **b** hospitalizations per 100 000 persons, by country of birth for countries with >10 000 residents in Norway (N=5 256 143). Models shown: 1) Unadjusted. 2) Base-model (age, sex, municipality of residence). 3) Base-model + socioeconomic factors (occupation, household crowding, education, household income). 4) Base-model + medical risk (14 diagnosis-groups). 5) Full model (age, sex, municipality of residence, occupation, household crowding, education, household income, medical risk). Bars indicate robust standard errors. BA-XK-HR-ME-RS-SI: Bosnia and Herzegovina, Kosovo, Croatia, Montenegro, Serbia and Slovenia.

Various sensitivity analyses were performed, including the number of people in the household in addition to the household crowding variable, and use of logistic regression models instead of linear. Main results were robust to these sensitivity analyses.

## DISCUSSION

Although factors related to social inequality were risk factors for infection and/or hospitalization with COVID-19 among both foreign-born and Norwegian-born persons, these factors only partially explained the differences across country backgrounds. The most obvious explanation for the high hospitalization rates in many of the groups is a correspondingly high infection rate (detected and undetected) (1). This is supported by high test positivity rates in many of the same groups with high hospitalizations (1), indicating persistent and extensive undetected infections in the immigrant population. Hospitalizations with COVID-19 is likely the best indication of the true levels of infection since it is independent of test activity, assuming that hospital capacity is maintained, and patients have good access to hospitals.

Our findings are somewhat more modest than a UK study (5) which found that adjustment for socioeconomic and lifestyle factors and comorbidities attenuated excess hospitalisations by 33% for Blacks and 52% for Asians. This may reflect that there are relatively fewer disparities in the Norwegian setting with strong social welfare rights and universal healthcare for all residents, including all registered immigrants. Nevertheless, our findings are in keeping with existing research that indicates that socioeconomic factors only partially attenuate disparities with regard to COVID-19 among ethnic minorities (4, 7, 8).

Large cities often have large proportions of immigrants, and urban living could be a factor in the spread of COVID-19 (9). Oslo has the highest proportion of residents with immigrant background and the highest notification rates of COVID-19 in Norway. Our analyses show that municipality of residence had the highest explanatory power for both infection rates and hospitalizations, however foreign-born persons had higher rates both in and outside of Oslo (2).

Immigrants often work in service-based occupations with close contact to others. However, a Norwegian study (10) found that immigrants from Somalia, Pakistan, Iraq, Afghanistan and Turkey, working in occupations with high contact frequency, did not have higher infection rates than others with the same country of birth, but did have higher rates than Norwegian-born with the same occupation. Networks related to the immigrant group therefore seem to be more important than the occupational network in explaining increased infection rates.

In some immigrant groups there might be a tradition for closer family ties than what is common in Norway. The high infection rates we see among Norwegian-born with foreign-born parents may indicate that much of the infection occurs within social environments that are connected to the parents’ country of birth. Because the virus spreads exponentially if no measures are implemented, even small increases in infection risk within a group of the population can quickly become significant if the group, due to strict regulations during the pandemic, has limited its social contact to people within its own group. Infection tracing data indicate that much of the spread occurs within families, and we find that household crowding is associated with infection regardless of country background. However, this variable, at least as it is defined, explains little of the increased infection rates among immigrants. It may be that it is difficult to stop the spread of infection within households regardless of crowding. More knowledge is needed as to how household factors affect the spread of infection, such as household composition, number of residents, multi-generational living, and housing-type.

The data used in our analyses only capture formally registered information. There are several factors that indicate that our estimates should be interpreted with caution, however the main findings should still hold. Foreign-born persons were more likely to have data missing for several variables. This may have contributed to poor explanatory power. Systematic biases in seeking medical help may affect registration of diagnosis codes on which medical risk groups are based. If so, medical risk will be underestimated in this study. However, a large study from the UK found that disproportionately high COVID-19 deaths among ethnic minorities were only partially reduced after adjustment for medical risk conditions in addition to socioeconomic status (8).

The overrepresentations we observe among foreign-born and their children may be due to a combination of several factors that are difficult to capture. For example, differences in travel patterns, how well the test-trace-and-quarantine strategy works in different groups, how infection spreads within a social environment, and interactions between different factors. Language barriers, vulnerable working conditions, and concerns about loss of income for those without the right to sick pay can all be barriers to testing, quarantine and isolation. Persistently high infection rates in some districts or municipalities may have led to a situation where high workload creates delays in contact tracing, in turn resulting in chains of infection not being broken. Although much has been done to adapt, translate, and disseminate information to the immigrant population, information about regulations and advice is complicated and in constant change. This can be challenging for all, let alone for people who do not master the local language.

## CONCLUSION

Residents with foreign-background have, as a group, been disproportionately hit by COVID-19 in Norway. Adjustment for socioeconomic factors and medical risk attenuates the overrepresentation moderately, however the overall picture remains the same. The data available or variable definitions may not have fully captured the effects and interactions of these factors, and future studies should aim to unravel this further.

## Data Availability

The datasets analysed in the current study are not publicly available due to privacy laws. Individual level data for research are generally available within Norway upon application conforming with strict regulations and procedures.

