## Supplemental methods for "SARS-CoV-2 infections and hospitalizations among immigrants in Norway – significance of occupation, household crowding, education, household income and medical risk. A nationwide register study"

**SUPPLEMENTAL MATERIAL**

**Supplemental Methods**

*Data sources*

The BeredtC19 Register is a national emergency preparedness register established to monitor infection and the use of health services in Norway during the coronavirus disease 2019 (COVID-19) pandemic (https://www.fhi.no/en/id/infectious-diseases/coronavirus/emergency-preparedness-register-for-covid-19/). BeredtC19 consists of individual-level data, covering the entire Norwegian population and includes information on country of birth. The register receives daily updates including from the Norwegian Surveillance System for Communicable Diseases (MSIS) and laboratory database (containing all tests for SARS-CoV-2 and test results), and the Norwegian Patient Registry (NPR). Data from the different national registries have been linked using the unique, personal identifier given to all Norwegian residents at birth or upon immigration. The BeredtC19 data used for this study originated from the Norwegian Surveillance System for Communicable Diseases and laboratory database (all polymerase chain reaction (PCR) tests with test results for SARS-COV-2), Norwegian Patient Registry and Norwegian Registry for Primary Health Care (hospital admissions, medical risk group), National Population Register (demographics, municipality of residence), Employer and Employee Register (occupation), and Statistics Norway (household crowding, education, household income).

*Study population*

The analyses include persons with a birth number in the National Population Register and who were residing in Norway as of 1 March 2020. Persons who have died or emigrated after 1 March are thus also included. We include the period from 15 June 2020 to 31 March 2021, excluding the first-wave due to limited test capacity and restrictive test criteria.

The sample consists of 5 494 669 people, of which 912 043 are registered with country of birth outside of Norway, and 199 518 are registered with both parents born outside of Norway. For several analyses we are primarily concerned with comparing people from the 25 largest immigrant groups in Norway with people born in Norway, and for these we have excluded people born in other countries from the analyses, giving a sample of 5 256 143 people in these analyses.

*Variable definitions*

Outcome variables: 1) A laboratory-confirmed infection (polymerase chain reaction test) with SARS-CoV-2 in MSIS, and 2) hospitalization with COVID-19 (NPR and MSIS). A COVID-19-related hospitalization is defined as a person who has tested positive for SARS-CoV-2 and has been hospitalized (24-hour stay) during the period 2 days before and 14 days after the positive test. We have also analysed whether a person has been tested for SARS-CoV-2 (laboratory database) and the proportion of the tested persons who have tested positive.

Country of birth**:** From the National Population Register. Foreign-born persons include all persons born outside of Norway, including those born abroad by Norwegian-born parents (a small number). Country of birth cannot be determined for persons who are not registered in the Population Register, and there are N=329 195 registered persons where country of birth is not stated, mostly older persons. In this study, we assume that these people were born in Norway.

For the overall analyses we operate with three categories i) Foreign-born, ii) Norwegian-born with two foreign-born parents (includes Norwegian-born with a foreign-born parent for persons registered with only one parent), and iii) Norwegian-born with one or two Norwegian-born parents. In these analyses, we define the land background of the Norwegian-born according to parents' country of birth (mother's if the parents were born in two different countries).

For all other analyses we include only foreign-born persons from the 25 countries / areas with at least 10 000 residents in Norway. Because the country of birth in the Population Register is not entirely reliable with regard to persons born in the former Yugoslavia, we have merged persons with these countries of birth in the category BA-XK-HR-ME-RS-SI: The name of the category refers to ISO 3166-1 alpha-2 codes, and are the country codes for Bosnia and Herzegovina, Kosovo, Croatia, Montenegro, Serbia and Slovenia respectively. Here we compare outcomes for persons born in each of the 25 countries with outcomes for persons born in Norway (where the latter group also includes persons born in Norway to foreign-born parents).

Explanatory variables

For adjusted models, explanatory variables are demographics (age, sex, municipality of residence), occupation, household crowding (crowding), education, household income, and medical risk group for severe COVID-19. All data are individual-level, or household level for crowding and income. Persons with missing data for any of the explanatory variables were coded into a separate residual category for that variable, thus keeping the same sample for all analyses.

Demographics**:** Age, sex and municipality of residence were obtained from the National Population Register. Municipality of residence was missing for N=95 343 (1.7%).

Occupation**:** From the Employer and Employee Register. For employees, we have information on occupation (2-digit STYRK-code) and industry (2-digit). We do not have information about occupation for self-employed persons.

Household crowding**:** A pre-defined indicator variable from Statistics Norway that takes the value 1 if the number of rooms in the dwelling is less than the number of residents (or 1 person lives in a 1 room dwelling), and if the number of square meters (P-area) is less than 25 $m^2$ per person. We have data on crowding up to and including 2019. Those who had a missing value for household crowding (N=405 642, 7.4%) were coded in a separate residual category.

Education**:** Highest completed education from Statistics Norway, registered as: Below upper secondary education; Upper secondary education; Tertiary vocational education; Higher education, short; Higher education, long; and undisclosed / no education. Tertiary vocational education level was included in these statistics only from 2016 and is therefore merged with Upper secondary education level in this report. We therefore use the following education categories in this study: 'Below upper secondary'; 'Upper secondary / vocational'; 'University / college, short'; 'University / college, long'; and 'Undisclosed / no education'. We have information on education up to and including 2019, and therefore we have coded people who are 25 years or younger per 1 March 2020 (N=1 606 768) into a separate category, to separate people who have not necessarily completed their education.

Household income**:** From Statistics Norway. Total household taxable and non-taxable income, minus taxes, divided by the number of consumption units in the household. The number of consumption units is calculated by using the 'modified' OECD scale or the EU scale, where the first adult is given a value of 1, any additional adult is given the value of 0.5, and each child under 17 years is given a value of 0.3. The number of consumption units in a household consisting of two adults and two children is thus 2.1, according to this method. After the household income has been adjusted according to family composition and size, it was divided into deciles. Data on income was available up to and including 2018, and people for whom we lack information about income (N=242 299, 4.4%) are placed in a separate residual category.

Medical risk group**:** A set of indicator variables for underlying diseases and health conditions that have been found to increase the risk of hospitalization for COVID-19 (1). The medical risk groups are defined on the basis of diagnostic codes registered in the primary and specialist health services back to 2017. There are 14 defined medical risk groups: organ transplantation; neurological diseases or muscle diseases that cause impaired coughing or lung function; chronic kidney disease or significant renal impairment; chronic liver disease or significant hepatic impairment; immunosuppressive therapy; diabetes; chronic lung disease (other than well-regulated asthma); obesity; haematological cancer during the last five years; other active cancer, ongoing or recently discontinued treatment for cancer; immune deficiency; chronic cardiovascular disease (with the exception of high blood pressure); stroke; dementia. See Nystad et al. (1) and Appendix 1 in (2) for further details and all included diagnosis codes.

*Statistical analyses*

We estimated the following linear probability model:

$$y_i = country_i \beta_k{}^{country} + controls_i^k \beta_k^{controls} + \varepsilon_i$$

Where $y_i$ is an indicator variable with the value 1 if individual $i$ has tested positive for SARS-CoV-2 / been hospitalizaed for COVID-19, and 0 otherwise. $country_i$ is a vector with indicator variables for birth country (reference category Norway), and $controls_i$ is a vector with covariates.

**Table A: Estimated models**

| *k* | **Model** | **Covariates** |
|---|---|---|
| **1** | Unadjusted | None |
| **2** | Demography | Age, sex |
| **3** | Residence | Age, sex, municipality of residence |
| **4** | Occupation | Age, sex, municipality of residence, occupation (2-sifret), industry (2-sifret) |
| **5** | Household crowding | Age, sex, municipality of residence, household crowding |
| **6** | Education | Age, sex, municipality of residence, highest completed education (Below upper secondary education, Upper secondary or Tertiary vocational education, short and long Higher education, undisclosed or no education, persons under 25 years old) |
| **7** | Income | Age, sex, municipality of residence, household income (deciles) |
| **8** | Medical risk group | Age, sex, municipality of residence, medical risk group (14 diagnosis groups) |
| **9** | All factors | Age, sex, municipality of residence, occupation, household crowding, education, household income, medical risk group |

Notes: Age was dummy-coded in 5-year categories for all models

For each of these specifications, we estimate a vector with coefficients for (groups of) countries of birth. These have an interpretation as expected difference in COVID-19 outcome between the different countries of birth and Norwegian-born persons, dependent on the covariates in the model.

For country of birth $countryC$:

$$\beta_k^{countryC} = E\left[y \middle| country = countryC, \overline{controls_k}\right] - E[y|country = Norway, \overline{controls_k}]$$

The estimates thus give the deviation in percentage points in relation to the mean of the reference group (typically Norwegian-born). To assess relative changes, not only absolute ones, we have divided each of the estimates by the mean incidence of COVID-19 for Norwegian-born persons, $\bar{y}_{Norway}$.

$$\tilde{\beta_k}^{country} = \frac{\beta_k^{country}}{\bar{y}_{Norway}}$$

$\tilde{\beta_k}^{country}$

thus has the interpretation as the percentage difference in expected infection between countries of birth (adjusted for the given observed differences).

The statistical programs Stata 16 and R were used for the analyses. Linear regressions (OLS) with robust standard error have been used to take into account that the residuals are not normally distributed for binary outcomes.
